# A comprehensive analysis of recovered COVID-19 patients and dynamic trend in antibodies over 3 months using ELISA and CLIA methods

**DOI:** 10.1101/2020.08.31.20184838

**Authors:** Puya Dehgani-Mobaraki, Asiya Kamber Zaidi, Alessandro Floridi, Alessandro Lepri, Emanuela Floridi, Alessia Gherardi, Enrico Bernini-Carri, Eleonora D’urzo, Massoud Dehgani-Mobaraki

## Abstract

**Background:** Since the Coronavirus disease-2019 outbreak, most studies have focused on etiopathogenic aspects and treatment strategies. Acquired immunity still remains a dilemma. The aim of our study included a comprehensive analysis of patient characteristics, evaluation of antibody response, and its trend over a period of three months in recovered patients.

**Methods:** Monocentric investigator-initiated pilot longitudinal observational study conducted by the Association Naso Sano, on a cohort of 30 COVID recovered patients based in the Umbria region, followed up from April to June 2020 for baseline blood counts, IgM and IgG trends using two different serological assays-ELISA and CLIA. The demographics, blood group, co-morbidities and treatment modalities were recorded from each patient along with an analysis of clinical profile, dates concerning symptom onset, first positive and two consecutive negative swabs using an online questionnaire followed by serological testing. Descriptive and Bivariate (Pearson correlation coefficient) statistics were conducted to detect statistically significant correlations.

**Findings:** The study involved 30 patients with a M:F ratio of 0.57 and a distribution of mild (67%), moderate (30%) and critical (3%). Majority of the patients were healthcare workers (40%) and the mean viral shedding duration was 20.13 ± 6.17 days. The IgG levels offered long-standing protection as long as 3 months in some cases. A statistically significant, directly proportional correlation (Pearson) exists between ELISA and CLIA values for IgM. Some patients also expressed titers lower than the detection threshold and therefore a positive RT-PCR test does not necessarily guarantee a high IgG response in the recovery period.

**Interpretation:** The data presented in our study provides a relative long-term analysis and possible explanation regarding the protection developed by patients recovered from COVID-19.

## Introduction

Since the outbreak of the Coronavirus disease 2019 (COVID-19), many studies have focused on the etiopathogenic aspects and treatment modalities. Since March 2020 different lockdown strategies have been adopted by majority of nations, while a few of them preferred a more flexible attitude by seeking the so-called herd immunity. This, however, did not mitigate the viral spread which currently has affected more than 10 million people worldwide and has caused more than 500 thousand deaths. Acquired immunity has become a relatively new dilemma for the recovered COVID-19 patients. Essentially limited to surveillance and epidemiological purposes, a precise assessment of the immune response and its duration would be beneficial. Our study aimed at a comprehensive analysis of patient characteristics, evaluation of their antibody response, and its persistence over a period of three months in a cohort of COVID-19 recovered patients.

## Methods

### Study design

A monocentric investigator-initiated pilot longitudinal observational study was conducted by the Association Naso Sano. This study was approved by the Research ethics committee of Association Naso Sano [Document number: ANS-2020/001]. The manuscript follows the CONSORT statement for the reporting of cohort studies.

### Participants

A cohort of 33 patients based in the Umbria region, who had tested positive for SARS-Cov-2 in March 2020 were included in the study. These patients were defined positive using the real-time PCR detection method from nasopharyngeal and pharyngeal swab specimens. Case definitions were based on the WHO guidelines. [1] Post recovery they were invited for serial serological tests by one of the authors, P.D.M, who had tested positive earlier this year and is also currently a part of the study sample.[2] Out of the 33 patients enrolled, three were excluded from the study: a 3 months old infant in view of negative consent from the parents for blood sampling and 2 patients who did not report for consecutive tests, making a final sample size of 30 patients.

### Data Collection

The participants of the study were followed up from April to June 2020 for baseline blood counts and trends for IgM and IgG using two different serological assays; ELISA and CLIA. The demographic characteristics, epidemiological data, blood group, co-morbidities, and treatment modalities undertaken were recorded from each patient. Also, the clinical profile, dates concerning symptom onset, first positive swab (S1), two consecutive negative swabs (S2, S3) were also recorded using an online 25-item based questionnaire. These patients were then invited for serologic tests (B1, B2), spaced minimum two weeks apart, using two different methods ELISA and CLIA (Figure 1). All patients were enrolled in the clinical registry and a written informed consent was obtained for voluntary participation from each patient. Local data protection policy was followed in order to record data securely.

**Figure 1:**
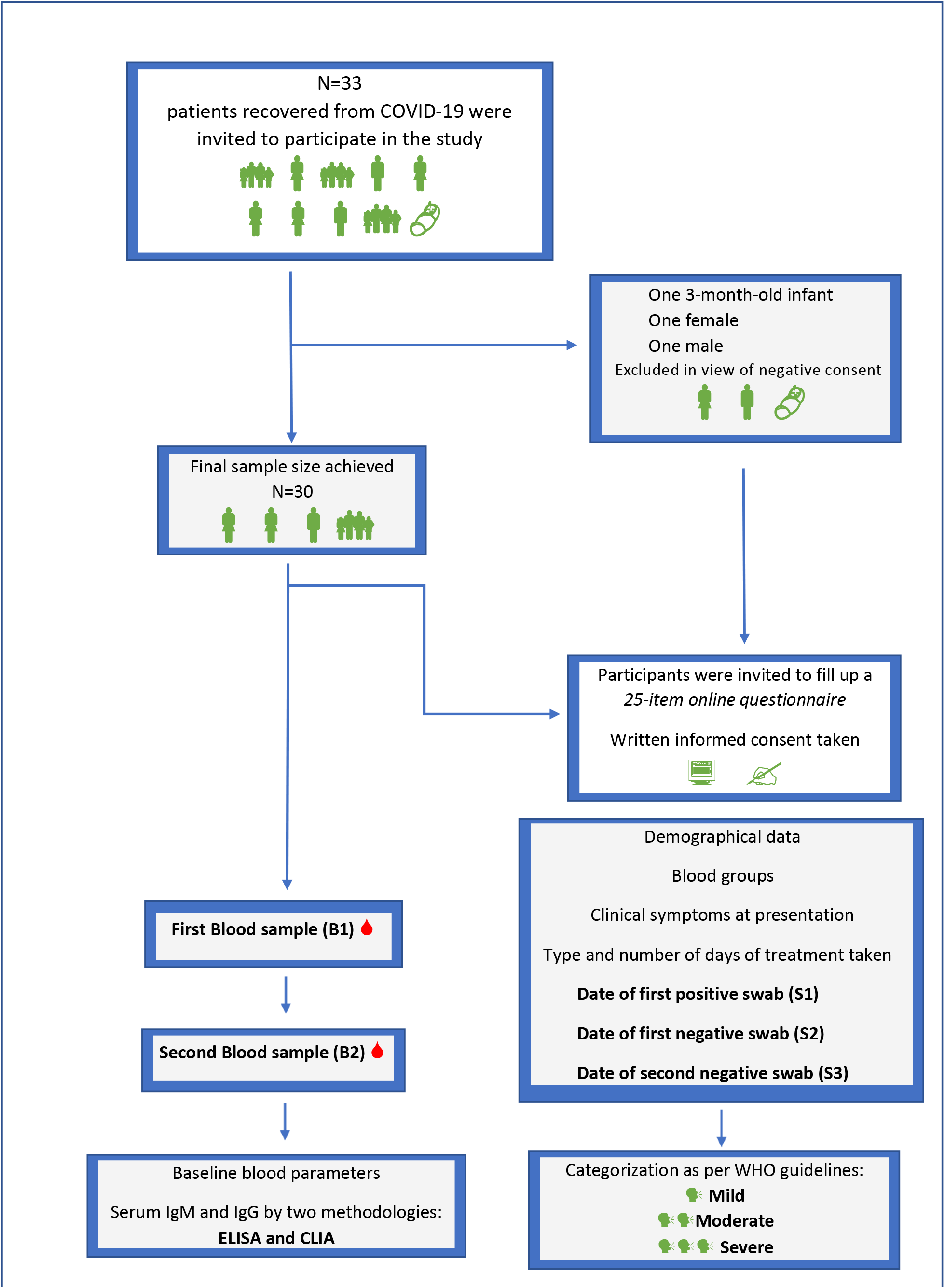
Study design flow diagram.

### Analytical systems used in our study

a. The NovaLisa^®^ SARS-CoV-2 (COVID-19) IgM/IgG ELISA assay: (NovaTec Immundiagnostica GmbH Waldstraße 23 A6, 63128 Dietzenbach, Germany). Results were reported in NovaTec Units [NTU]. Sample (mean) absorbance value × 10/ cut off = [NovaTec Units = NTU].[3]
b. The MAGLUMI^®^ 2019-nCoV lgM/lgG chemiluminescent analytical system (CLIA) Assay: (New Industries Biomedical Engineering Co., Ltd [Snibe], Shenzhen, China). Results were reported as measured chemiluminescence values divided by the cut off (absorbance/cutoff, S/CO): S/CO >1 was defined as positive and S/CO≤1 as negative.[4] All tests were performed under strict biosafety conditions in the same lab.

### Statistical Analysis

Statistical analysis was carried out using SPSS [version 25, IBM] and statistical functions were used to plot co-relations between variables. The descriptive statistics (Mean [M], Standard deviation [SD], Min., and Max.) were conducted to explore the features of the sample. Bivariate statistics (Pearson correlation coefficient) were conducted to detect a statistically significant correlation between dependent variables: IgM ELISA with IgM CLIA levels and IgG ELISA with IgG CLIA levels.

## Results

### Participants’ description

Between April and June 2020, we invited 33 patients recovered from COVID-19 to participate in our study, and out of these, three participants were excluded. The reasons for exclusion were 3-month-old infant (n=1) and negative consent for consecutive tests (n=2). Of the 30 patients included in our study, 11 (37%) were male and 19 (63%) females. The overall mean age was 47.16 ± 17.81 years (min=21; max=81), with 54.36 ± 18.05 years for males and 43 ± 16.7 years for females. The study sample was diverse and included family clusters along with a pair of twin females, one of them was in her post-partum state, and an unrelated antenatal pregnant female. Blood groups were recorded and divided based on sex and clinical severity [5] as mild (20 cases, 66.6%), moderate (9 cases, 30%), and critical (1 case, 3.33%). The percentage of females was higher than that of males in the mild (60% female and 40% male) and moderate (77.77 % female and 22.22% male) groups, while the critical group involved only one male patient (3.33 %) (Figure 2a).

**Figure 2:**
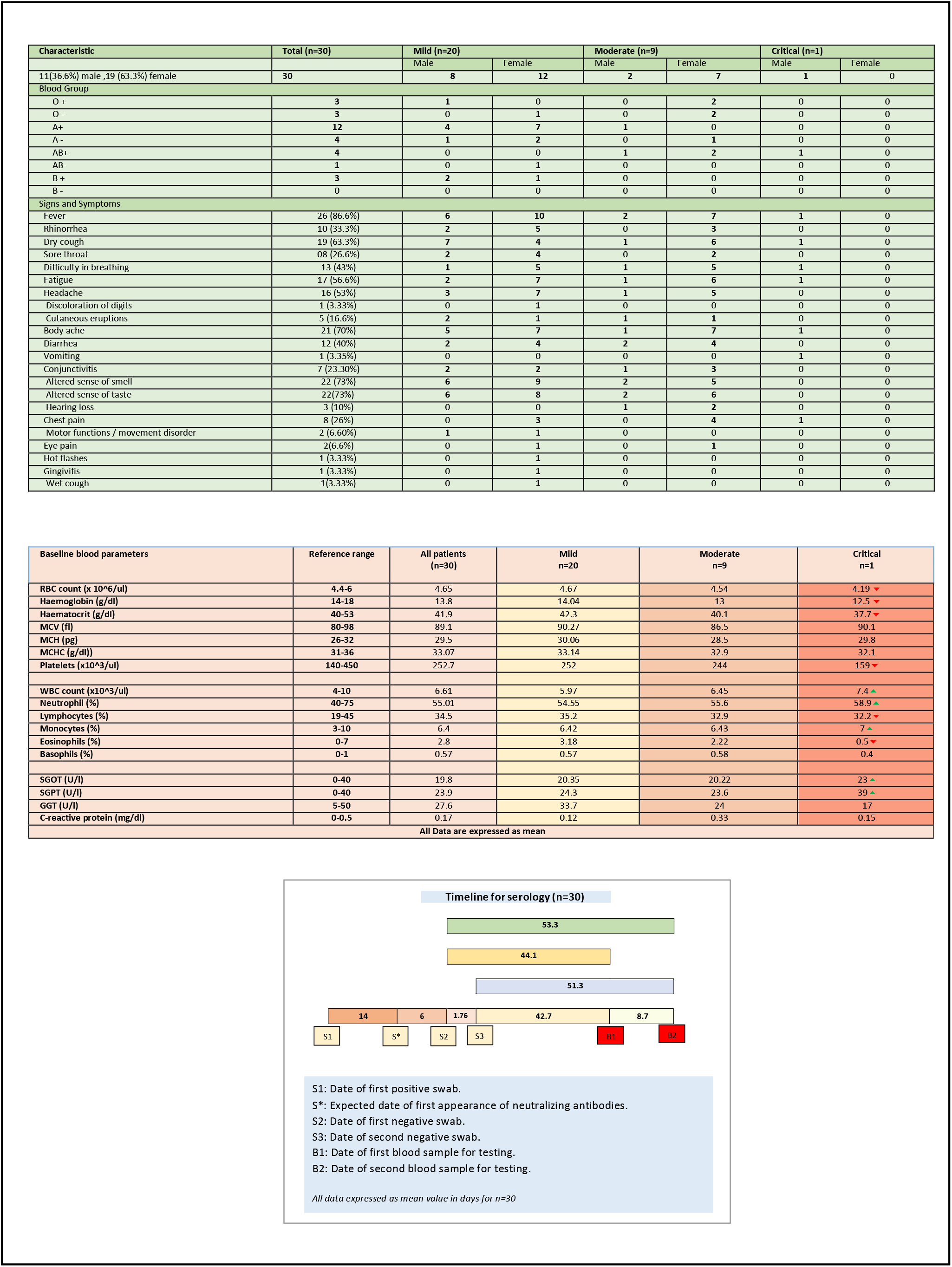
Table showing (a) demographics, blood group, sign and symptoms; (b) baseline blood parameters; (c)Timeline for consecutive swabs, blood tests and duration between them expressed as mean for all patients (n=30)

Majority of the patients were healthcare workers (12 patients, 40%) followed by general employees (7 patients, 23.3%), retired (5 patients, 16.66 %), businessman/woman (2 patients, 6.66 %), students (2 patients, 6.66 %), homemakers (2 patients, 6.66 %). The duration for viral shedding (number of days between S1 and S2) was expressed as a mean of 20.13 ± 6.17 days with a median of 21 days.

Comorbidities were present in nearly half of patients, with hypertension being the most common comorbidity (n=6, 20 %), followed by diabetes (n=2, 6.66 %) and coronary heart disease (n=2, 6.66%). The number of patients with asthmatic and allergic tendencies was n=5, 16.6%. The most common symptoms on admission were fever (n=26, 86.66%), followed by altered sense of smell (n=22, 73%) and taste (n=22, 73%), muscle pain (n=21, 70%) and sore throat (n=19, 63.3%). Out of the 33 patients, n = 13 (43.3 %) patients received antibiotics and n= 1 (3.33 %) needed hospitalization. (Figure 2a)

A complete analysis of blood parameters, B1 and B2 for each of mild, moderate and critical groups, was done (Figure 2b and 2c).

### The effect of antibody trends over time

Blood samples were collected 44 days after S2 (mean value) and 53 days after S2 (mean value) and named B1 and B2 respectively. Trends for IgM and IgG were assessed from B1 to B2 by both ELISA and CLIA methods for mild, moderate, and critical groups (Figure 3, 4).

**Figure 3:**
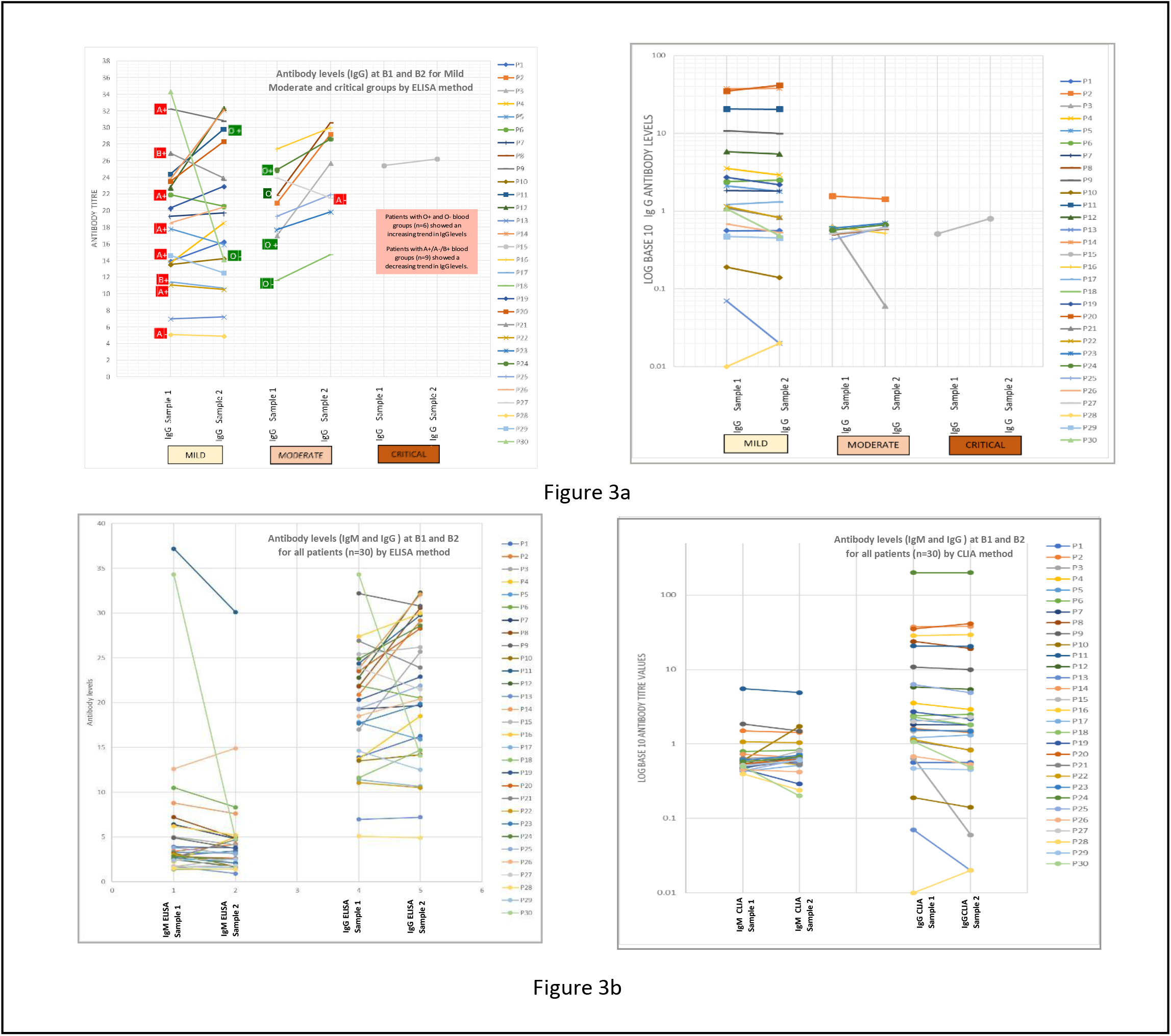
(a) Effect of blood groups on IgG levels for mild, moderate and critical groups by ELISA and CLIA methods at B1 and B2; (b) Antibody trend (IgM and IgG) for each patient over time.

**Figure 4:**
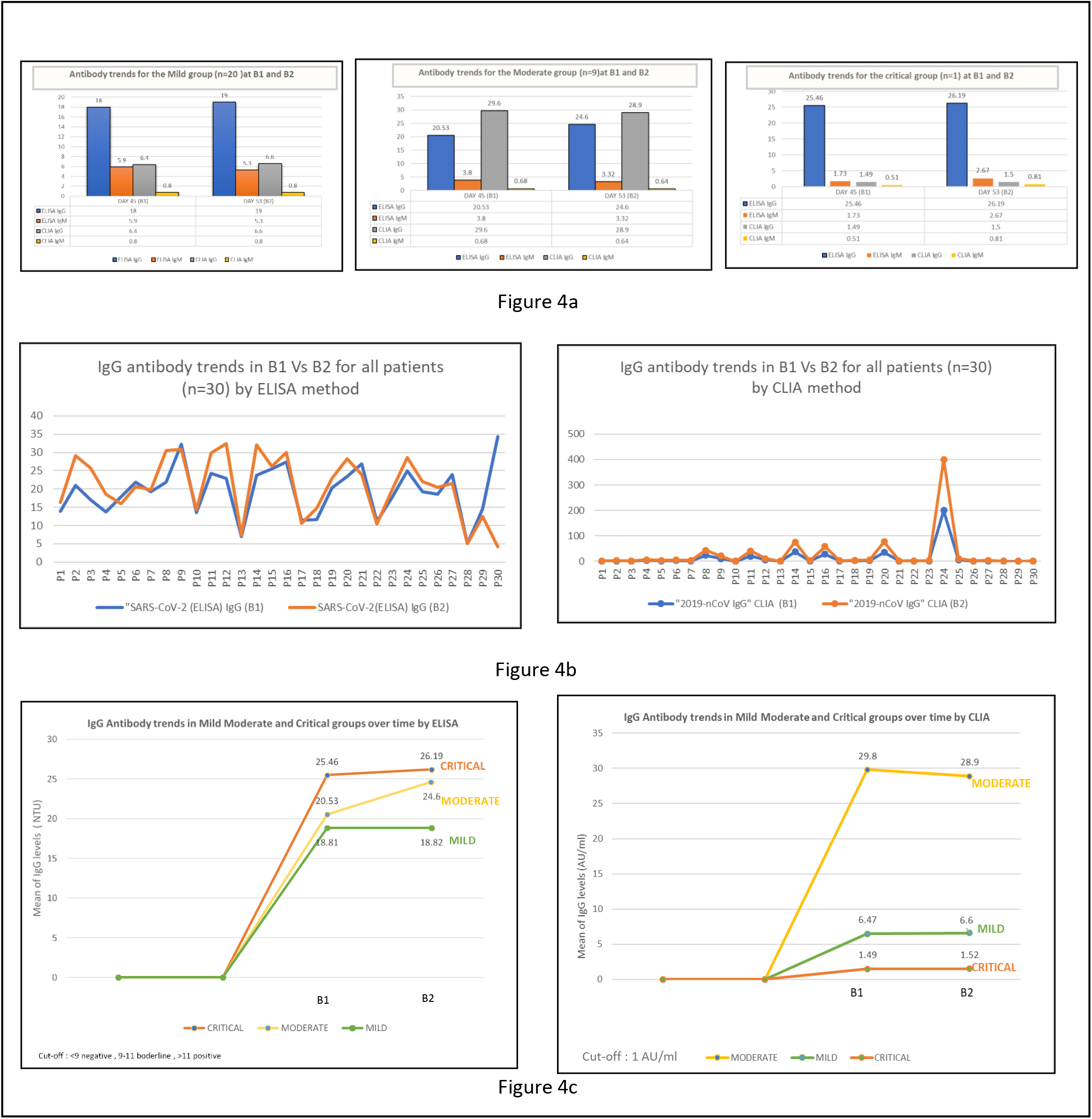
(a) Mean antibody titres (IgM and IgG) for mild (n=20), moderate(n=9) and critical(n=1) groups at B1 and B2 by ELISA and CLIA methods; (b) Overall IgG trend for all patients (n=30) at B1 and B2 by ELISA and CLIA methods; (c) Mean IgG values for mild, moderate and critical groups at B1 and B2 by ELISA and CLIA methods.

**Figure 5:**
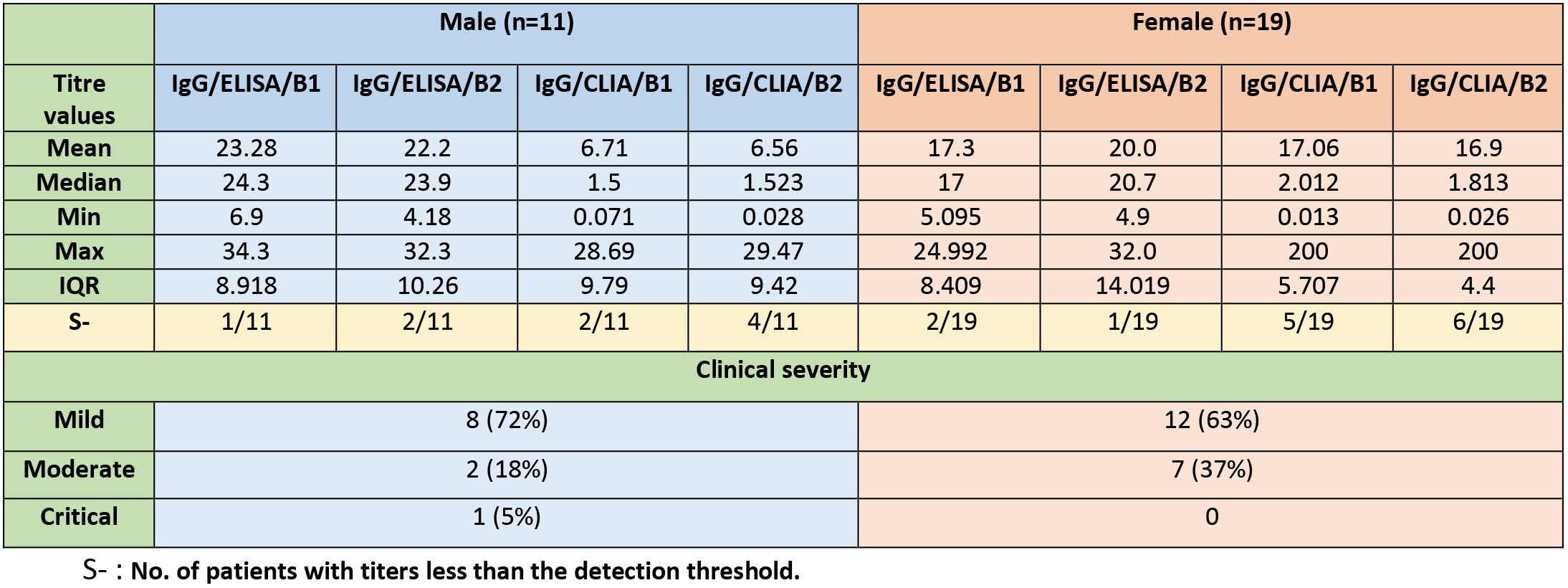
Gender based distribution of IgG titre kinetics by ELISA and CLIA methods, seronegative proportions and clinical severity.

### Pearson’s correlation between CLIA and ELISA assay performance

We found a positive correlation r = 0.870, n = 30, p<.000 between the IgM antibodies detected by the ELISA and CLIA methods at B1 and a positive correlation r = 0.775, n = 30, p<.000 between the IgM antibodies detected by the ELISA and CLIA methods at B2.

One-way and multivariate ANOVAs were conducted to compare the effect of the independent variables extracted from the socio-anagraphic and epidemiological questionnaire on the levels of IgM and IgG detected by ELISA and CLIA methods but since the sample size was small (n=30) they have not been described in this study. A sample size with a larger number of patients would be required for a significant statistical result. The statistically ideal sample size was determined by performing a *priori power analysis* [6] taking into account the research design including an independent 2-level variable (B1 and B2). Of these 2 independent variables, the B1 condition (i.e. the first blood sample) is used as control group compared to the others. In practice the same subjects are measured longitudinally and in our case 2 times. This type is called “repeated measures design”, as each subject is measured several times for a dependent variable. [7] This typology is typical of *within* variables. The statistical calculation made considered a minimum threshold of the power of the study (power = .80), a significance of the effects at p = .05 and an average size of the effect in the population. The calculation determined an ideal sample of 128 subjects.

## Discussion

As the number of patients testing positive for SARS-Cov-2 continues to rise, a significant number are on their way to recovery. A guarantee of the efficacy and duration of acquired immunity following this infection now requires our maximum attention. At present, there are not many studies on the duration of this immunity nor are there scientific evidences concerning the relationship between the severity of pathology and the antibody titers. Our study has highlighted the development and persistence of IgG antibodies with an increase in trend over 2 months after recovery. The mean duration of viral shedding, expressed as the days between the first positive swab (S1) and the first negative swab (S2), was 20.13 days (min=11 days, max=43 days, median =19 days, IQR= 6 days). A recent study by Quan-Xin Long et al. [8] showed that asymptomatic patients had a significantly longer duration of viral shedding than the symptomatic with a log-rank P=0.028, while our results demonstrated that viral shedding duration varies significantly and might also depend on the severity of disease. In an evaluation of patients recovering from severe Covid-19, Zhou et al. found a median viral shedding duration of 31 days (range, 18-48 days). [9] It appears that the severity of infection plays a role in the IgG trends over time based on the methodology adopted. In our study, the ELISA method demonstrated an increasing trend for all three groups (mild, moderate, and severe) with the highest level for the critical group followed by the moderate group and the least values for the mild group. The CLIA method demonstrated an increasing trend only for the mild and critical groups.

Neutralizing antibodies provide specific immune defense against viral infections. Recent studies have revealed that the Sars-Cov-2 neutralizing antibodies could be detected in patients from day 10-15 after the onset of the disease [10], in some cases, IgG would appear even earlier within 7 days by ELISA [11] and within 14 days by CLIA [12], with individual differences in dynamics both in terms of age and severity of the disease.

Neutralizing antibodies in asymptomatic patients seem to decrease within 2–3 months post-infection [6], however a recent Cochrane Review reported very few studies with data regarding antibodies duration beyond 35 days in symptomatic patients. [13]

Our study demonstrated a persistent IgG antibody response in 20 out of 30 patients by the ELISA method and in 25 by the CLIA method at B2, resulting in a positive immune response for a duration of minimum 34 days and a maximum of 67 days after the first negative swab (S2).

Since the mean between S1 and S2 was 20.13 days, the duration of immunity offered by the IgG might be longer. To understand this, we could assume that, if we calculate the day when the neutralizing antibodies could be detected (S*), meaning 14 days after S1 and 6 days before S2, then the duration of persistence of IgG would range from a minimum of 40 and a maximum of 73 days. This finding suggests that IgG levels persist offering long-standing protection for close to 3 months in some cases. (Figure 3)

When the patients were divided into three groups based on their age, young (15-39 years), middle-aged (40-59 years), and elderly (60-85), it was observed that middle-aged patients were more likely to induce higher titers of IgG than young and elderly patients.

Regarding the behavior of the two methodologies (ELISA and CLIA), it was observed that there exists a statistically significant and directly proportional correlation (Pearson) only between the IgM values and similar findings were reported by Padoan et al. [14]

It was also observed that some patients did not have antibodies or had titers lower than the detection threshold post-exposure. It is noteworthy that a positive RT-PCR test does not necessarily guarantee a high IgG antibody response in the recovery period. In our study, while all patients (n=30) had a positive swab, not all patients had an IgG response above the detection threshold levels. For B1, by the ELISA method, 2 patients had a lower than the cut-off IgG antibody levels (negative) and by the CLIA method, this number was 7. For B2, by the ELISA method, 3 patients had negative results and 2 were borderline, while 10 were negative by the CLIA method.

This could be due to protection through an alternative pathway of immunity such as cell mediated immunity. [15] The trend of antibodies needs to be studied over a longer period in order to come to a significant conclusion.

The strengths of our study include its monocentric, investigator-initiated, and unblinded study design with a diverse sample quality involving multiple family clusters from the same region. The serological tests were conducted by the same laboratory and a single team of doctors analyzed the data using the same clinical approach. Daily online meetings ensured a complete assessment of the patients’ data and re-checks wherever inconsistencies were encountered.

The patient population included mild, moderate and only one critical case, and hence only one out of the 30 patients needed hospitalization.

The limitations of our study included its small sample size (30) and analysis of only the antibody titers which might not be sufficient to assess the overall immunity of the patient, as some studies have proven that both, the lymphoid and myeloid immunity have a role to play in offering protection against future infection. [15] While the humoral immunity comprises the action of antibodies developed against the infection; the Cell-mediated immunity involves the action of T cells.

Yet another limitation could be the fact that the relationship of the measured antibodies with that of neutralizing activity against SARS-CoV-2 was not evaluated. The neutralizing antibodies are detected by the Plaque reduction neutralization test (PRNT), which requires a live virus and a biosafety level of 3. Few studies, however, have demonstrated the antibodies that target different domains of S protein, including S1, RBD, and S2 might contribute to “virus neutralization”. [16, 17] Inhomogeneity of the sample concerning blood groups and clinical severity could be attributed to the study being cross-sectional and observational. Moreover, most of these assays in the market are based on studies conducted on severe hospitalized patients [13] creating a “spectrum bias” and so these might not be generalized for patients with mild symptoms.

## Conclusion

The evaluation of antibody levels represents a cornerstone in the classification of a disease. The data presented in our study provides a relatively long-term analysis and possible explanation regarding the protection developed by patients recovered from COVID-19. At present, with a rapidly evolving pandemic, the reassurance for a protective immune response post-recovery is essential.

## Data Availability

All the data of the study will be available for review.

## Acknowledgments

The authors would like to acknowledge all the participants involved in the study. The laboratory (CRABION Pvt. Ltd.) and the phlebotomists provided in-kind support, in the form of sampling, equipment, and consumables for the evaluation, but had no role in directing the study or influencing the study outcomes. A special acknowledgement goes to my daughter and her magnificent smile that gave me the strength to fight and remain calm during my battle to fight my Covid-19 disease.

## Research funding

None declared.

## Author contributions

All the authors have accepted responsibility for the content of this manuscript and approved its submission.

## Consent

Informed consent was obtained from all the individuals enrolled in this study.

## Ethical approval

The study has been cleared by the local Ethical Committee of *Association Naso Sano* (Document number: ANS-2020/001).

